# Amplitude setting and dopamine response of finger tapping and gait are related in Parkinson’s disease

**DOI:** 10.1101/2021.05.17.21257331

**Authors:** Hafsa Bareen Syeda, Aliyah Glover, Lakshmi Pillai, Aaron S. Kemp, Horace Spencer, Mitesh Lotia, Linda Larson-Prior, Tuhin Virmani

## Abstract

**Objective:** Movement amplitude setting is affected early in Parkinson’s disease (PD), clinically manifesting as bradykinesia. Our objective was to determine if amplitude setting of upper limb bimanual movements and bipedal gait are similarly modulated in PD.

**Methods:** 27 PD and 24 control participants were enrolled. Participants performed a bimanual anti-phase finger tapping task wearing gloves with joint angular sensors, and an instrumented gait assessment. Motor load was varied by asking participants to perform movements at a normal and fast pace. PD participants were evaluated OFF (PD-OFF) and ON (PD-ON) levodopa.

**Results:** PD-OFF participants had shorter tap interval, smaller tap amplitude, and greater tap amplitude variability than controls in the more affected hands (all p<0.05). Tap amplitude and stride length (p=0.030), and tap frequency and gait cadence (p=0.011) were correlated in PD-OFF. Tap frequency and amplitude were also correlated with motor UPDRS (p<0.005) and bradykinesia motor (p<0.05) and ADL (p<0.005) UPDRS subscales. Levodopa improved mean tap amplitude and stride length during fast tapping in PD participants.

**Conclusion:** In PD-OFF, mean finger tapping amplitude and gait stride length were correlated and showed similar dopaminergic response.

**Significance:** Future studies manipulating upper limb amplitude could help provide greater understanding of the networks responsible for amplitude setting in PD.

## Introduction

Bradykinesia is defined as a slowing of movement in addition to a tapering amplitude with repetitive movements. By contrast hypokinesia refers to slowing of movement alone. Limb bradykinesia, based on the UK brain bank (Hughes et al. 1992) and MDS criteria (Postuma et al. 2015), is required for a clinical diagnosis of Parkinson’s disease (PD). Upper limb bradykinesia can lead to significant functional decline in tasks requiring bimanual coordination such as cutting meat, buttoning clothes and shampooing one’s hair. While these tasks may not have the same rhythmicity of movement that gait does, they are still learned complex patterned automatic movements that do not require significant thought during their performance.

Studies to address deficits in upper limb movements have used multiple paradigms including bimanual in-phase (limbs moving together in the same direction) and out-phase (limbs moving in opposite directions) actions (Almeida et al. 2002; Ponsen et al. 2006). Different movement types have been studied by different groups; including proximal arm displacements towards and away from the body (Almeida et al. 2002), pronation/supination, wrist flexion-extension (Almeida and Brown 2013), circular drawing movements (Ponsen et al. 2006) and even more complex tasks such as moving one’s hand to the mouth (Corona et al. 2018). A wide array of technologies have also been employed to capture upper limb movements including mechanical sliders (Almeida et al. 2002), tablets (Nieuwboer et al. 2009), MIDI keyboards (Koop et al. 2008), angular sensors (Vercruysse et al. 2012a), and 3D motion capture (Delval et al. 2017). Upper limb bimanual coordination deficits have been shown to occur early in the PD disease course (Ponsen et al. 2006). PD participants have more difficulty with anti-phase movements, showing greater variability and more frequent switching to an in-phase pattern than do healthy controls (Almeida et al. 2002; Johnson et al. 1998).

Studies comparing upper and lower limb function in the same participants are limited. Using a task instructing participants to move their finger between two dots a fixed distance apart, Williams et al. (Williams et al. 2013) calculated a phase coordination index using the time to move between the two dots. For this upper limb task, performed in the OFF-levodopa condition only, they set upper limb movement speeds using a metronome timed to gait cadence, and found a correlation with the phase coordination index for gait at the participants preferred speed. Delval and colleagues (Delval et al. 2016) evaluated early PD participants in the OFF-levodopa condition using constrained tapping movements of the fingers and feet to set metronome frequencies while gait speed was not constrained. They found that freezing and festination episodes in finger and foot tapping were present before gait freezing developed in some, suggesting that a break-down in repetitive upper and lower limb movements may precede the break-down in more complex movements such as gait. The kinematic relationships between finger tapping, foot tapping and gait were not explored. Barbe and colleagues (Barbe et al. 2014) primarily compared variability in finger tapping and variability in gait stride length to occurrences of freezing events independently for upper limb and gait. They report that smaller step length was associated with the occurrence and duration of gait freezing episodes, while the variability in upper limb movements was related with the occurrence and duration of upper limb freezes. While measurements of finger tapping amplitude and gait stride were performed in the OFF-state and ON-state, they did not directly compare mean or variability in finger tapping amplitude and stride length in either condition.

No studies have been conducted to our knowledge in people with Parkinson’s disease to determine whether the amplitude setting of upper limb movements and gait show similar deficits. As upper body parkinsonism is a more common early disease manifestation than gait deficits, evaluation of pathways that are disrupted at the earliest stages of PD using finger tapping paradigms could provide earlier insight into putative targets for pharmaceutical intervention or neuromodulation. Current methods to study networks involved in gait are limited in a Magnetic Resonance Imaging (MRI) scanner as studies have to be performed lying down with assessments using virtual gait environments with foot pedal movements (Shine et al. 2013). We hypothesized that both upper limb bimanual movements and gait movement amplitude would be similarly modulated in PD participants. If this were true then we would expect that finger tap amplitude would be reduced proportionally with gait stride length in PD participants. We also would expect movement amplitude to be similarly modulated in upper limb movements and gait during higher motor stress. To test this hypothesis, we had PD participants in the levodopa OFF-state and healthy age matched controls perform both bimanual anti-phase finger tapping wearing data gloves (with a sensor on the metacarpophalangeal joint), and walk on an instrumented gait mat. Both gait and finger tapping were performed at self-defined normal and fast speeds (to test differences in motor loading/stress). PD participants treated clinically with levodopa were subsequently assessed in the levodopa ON-state to determine if dopaminergic response was similar between the upper limb and gait tasks.

## Methods

### Standard Protocol Approvals, Registrations, and Participant Consents

Participants were recruited from the Movement Disorders Clinic at the University of Arkansas for Medical Sciences (UAMS). The study was approved by the UAMS institutional review board (UAMS IRB# 228861), written informed consent was obtained from all participants, and the study was conducted in accordance with the guidelines of the Declaration of Helsinki.

### Study population

Participants with PD based on UK brain bank diagnostic criteria (Hughes et al. 1992), and age-matched controls (controls) between the ages of 45-90 were enrolled. 51 participants were enrolled (24 controls, 27 PD) and analyzed. Exclusion criteria included inability to walk on the Zeno walkway, falls > 1/day, cognitive impairment sufficient to impair capacity for informed consent, diagnosis of a neurologic disorder (other than PD for the PD group), diagnosis of a psychiatric disorder other than those associated with PD, the use of anti-dopaminergic medications in the year prior to enrollment, chronic back, hip or knee pain that was not controlled, severe osteoarthritis, hip or knee replacement surgery or spine surgery in the last 12 months or complicated by persistent pain, and inability to complete questionnaires in English. A complete Unified Parkinson’s Disease Rating Scale (UPDRS), a Hoehn and Yahr staging score (H&Y), the Montreal Cognitive Assessment (MoCA)(Nasreddine et al. 2005), and the Hamilton depression (HAM-D)(Hamilton 1960) and anxiety (HAM-A)(Hamilton 1959) rating scales were also performed on all participants. UPDRS Bradykinesia motor (sum of UPDRS items 23-26 and 31) and activities of daily living (ADL) (sum of UPDRS items 9-12) subscores were calculated for PD participants.

### Disease asymmetry calculations

In PD participants, the side more affected (MA) by PD was calculated based on the ratio of the summated right/left scores for items 20-26 of the UPDRS corresponding to tremor, bradykinesia and rigidity, with a score > 1 indicating right-side more affected by PD. All data (other than Figure 1 which also shows the less affected (LA) side are reported for the more affected (MA) side.

**Figure 1:**
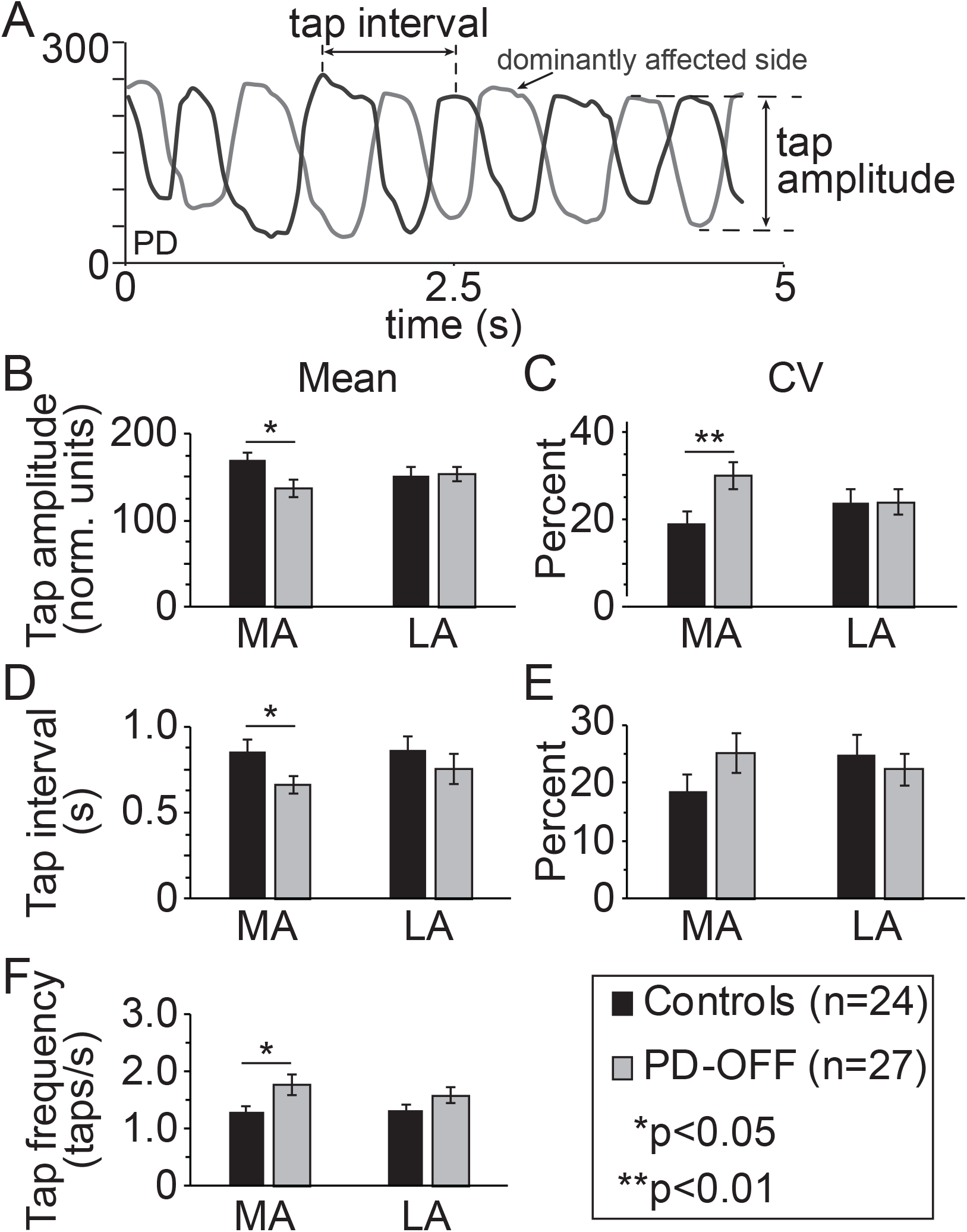
Spatiotemporal parameters of upper limb finger tapping. **(A)** Sample trace showing bimanual finger tapping output using the data glove in a participant with PD in the levodopa OFF state. Definitions of tap interval and tap amplitude are shown. Mean and variability (CV) in finger tap amplitude **(B, C)** finger tap interval **(D, E)**, and tap frequency **(F)** for the more affected (MA) and less affected (LA) hands are shown for controls and PD participants OFF levodopa (PD-OFF). Bar graphs are plotted as the mean ± sem.

### Upper limb dynamics

Participants were placed in a comfortable seated position at a desk with data gloves on (5DT Data Glove, Fifth Dimension Technologies Inc., Orlando, FL), instructed to rest the palms of their hands on the table and alternately tap the right index finger and left index finger on the table; i.e. while the right index finger was moving towards the table, the left index finger would be concurrently lifting off the table (out-of-phase tapping) and vice-versa. This task was chosen to simulate bipedal gait where the right and left leg alternately move forward in a patterned manner, with one leg advancing at a time. As the other leg remains planted for the majority of the gait cycle during which the other leg is moving forward, it is effectively moving backwards in relation to the center of gravity. This task was performed for 20 seconds at (1) a “comfortable” pace (normal speed) and (2) as fast as possible (fast speed). The normal speed task was performed to mimic normal speed of gait and was not constrained to a metronome as has been done in some prior studies (Stegemoller et al. 2009; Williams et al. 2013), as gait studies are also not constrained to a metronome unless auditory cuing tasks are being tested. Alternating tapping movements were confirmed by visual examination during the task. The fast speed tapping was performed to test the response of participants’ movement to a task requiring increased motor complexity or motor load. The same motor loading task was performed for gait by asking participants to walk at a fast speed.

Data was recorded using 5DT Glove Manager Software, which generates a CSV data file. The glove uses an 8-bit A/D convertor providing 256 intermediate positions between a flat and fisted hand. An intrinsic algorithm in the software adjusts the raw values collected to the maximum and minimum sensor angles, with the hand flat and fisted positions to account for the effect of varying hand sizes (Burdea and Coiffet 2017). To calculate tap interval and tap amplitude of right and left index finger from these sinusoidal recordings, a protocol using Python and Visual Basic for Applications (VBA) was developed. Finding peaks in a signal depends on distinguishing between actual peaks and noise /baseline changes. Considering the noisy signals, we decomposed the peak detection procedure into two parts: calculating an amplitude threshold for peak selection and peak/trough detection. For amplitude threshold selection, the average of normalized glove readings was found to be too high to optimize the peak detector to distinguish between actual peaks and noise. Upon further iterations, the absolute difference between frames/sets of 10 values and the absolute difference between individual values was calculated and averaged. The difference between the two averages produced the most accurate threshold for peak detection. Using this threshold, peaks and troughs were detected by the local maxima and local minima method (Yang et al. 2009) and automated in VBA for all participant files. Python Pandas and Numpy libraries were used for peak/trough finding. The signal was then plotted against time with marked peaks and troughs using Matplolib.

The difference between the value of the trough of the sinusoidal curve to the next peak was used to calculate the amplitude of each finger tap (see Figure 1A). This value is a normalized output with a range from 0-255 units based on the flat and fisted configuration of an individual’s hand and will be referred to in terms of normalized units. The mean and percent coefficient of variability (CV) for finger tap amplitude across each trial was calculated independently for each hand. Two timing variables were also calculated in addition to tap amplitude to compare and contrast any changes in amplitude measures. The tapping frequency was calculated as the number of taps divided by the 20s trial duration. The mean and CV intertap interval was calculated independently for each hand from the time interval between the peaks on the sinusoidal tapping traces for each finger tap (see figure 1A). As phase coupling between hands (and legs for gait) was not the primary interest of this study, the hands were analyzed independently - any phase changes that may have occurred during the tapping task were not considered in the analysis.

### Gait kinematics

Participants were instructed to walk at (1) a “comfortable” pace (normal speed) and (2) as fast as possible (fast speed), 8 lengths of a 20’x4’ instrumented gait mat, and data was collected and analyzed using the Protokinetics movement analysis software (PKMAS, Protokinetics, Haverton, PA). The two speeds were chosen to correspond to the normal and fast speed (to assess motor load) performed during finger tapping. The mean and CV for continuous gait stride length, and stride time as well as gait cadence (steps/minute), were extracted using the intrinsic algorithms of PKMAS. These variables were chosen as amplitude, timing and frequency gait variables to correspond to the finger tapping measures defined above.

### Dopaminergic response

All PD participants underwent UPDRS, upper limb dynamics and gait kinematic measurements in the morning in their effective levodopa OFF-state after withholding their Parkinson’s medications overnight as per prior protocols (PD-OFF)(Delval et al. 2016; Hausdorff et al. 2003; Williams et al. 2013). PD participants who were taking levodopa (n=23) as part of their clinical regimen were examined again 60 minutes after their regular morning dose of levodopa in order to determine if amplitude setting in the upper limb and gait both responded to levodopa (PD-ON). Participants were not excluded from participating if they were not clinically treated with levodopa as the majority of the analysis was performed in the levodopa OFF-state. OFF-state and ON-state assessments were performed to determine if any improvement in gait and upper limb dynamics with levodopa were the same.

### Statistical analysis

Statistical analysis was performed using SPSS 24 (IBM). Our primary interest was comparing mean and variability in amplitude measures in the upper limb (finger tap amplitude) and gait (stride length). Timing and frequency measures were also compared as a contrast to the amplitude measures. Linear regression analysis and Pearson’s correlation coefficients were used to compare objective spatiotemporal gait and finger tapping measures in the PD group. A post-hoc Benjamini-Hochberg adjustment was applied for the multiple comparisons performed in Table 3. A student’s t-test was used to compare the control and PD group differences. Secondary interest was to determine if amplitude setting in upper and lower limb movements were similarly regulated when exposed to a motor load through speeded movements, or dopamine repletion through measurement of levodopa responsiveness (OFF-ON dopaminergic medications). A paired student’s t-test was used to compare results in the OFF-state and ON-state in the subset of 21 PD participants who completed both assessments. Repeated measures analysis of variance (ANOVA) was utilized to calculate the combined group (control/PD) and speed of tapping (normal/fast speed) differences.

## Results

Age was well matched between the PD and control groups (PD 69.3±8.4 years, controls 66.5±8.1 years, p=0.228) but the gender distribution was opposite in the two groups (PD 29.6% female, controls 66.7% female, p=0.008) as most controls were spouses of the PD participants (Table 1). PD participants had a mean baseline MoCA score that was two points lower than controls (PD 26.1±3.4, controls 28.0±1.7, p=0.013) although the mean score in both cases remained in the normal range for the test (≥26) (Table 1). Depression and anxiety scores were also higher in the PD group (Table 1). Other features of the PD group are also noted in Table 1.

**Table 1:** Demographics

| Table 1: Demographics |  |  |  |
| --- | --- | --- | --- |
|  | HC<br>(n=24) | PD<br>(n=27) | p-value |
| Age at enrollment (years) | 66.5 ± 8.1 | 69.3 ± 8.4 | 0.228 |
| Gender (female/male) | 16/8 | 8/19 | **0.008** |
| Right-handed | 100% | 81% | 0.085 |
| OFF UPDRS Part III (motor) Score | 4.1 ± 3.4 | 24.4 ± 11.2 | **<0.001** |
| OFF Total UPDRS Score | 6.7 ± 4.5 | 40.1 ± 18.5 | **<0.001** |
| ON UPDRS Part III (motor) Score |  | 16.6 ± 10.3 (n=22) |  |
| ON Total UPDRS Score |  | 28.1 ± 17.4 (n=22) |  |
| Motor UPDRS (OFF-ON) |  | 7.8 ± 6.9 (n=22) |  |
| MOCA score | 28.0 ± 1.7 | 26.1 ± 3.4 | **0.013** |
| HAM-D | 2.9 ± 3.2 | 6.3 ± 4.7 | **0.004** |
| HAM-A | 2.7 ± 3.0 | 4.7 ± 3.6 | **0.018** |
| Right side more disease affected (MA) | - | 56% |  |
| PIGD phenotype at visit | - | 41% |  |
| Initial symptom (rest tremor/gait) | - | 48%/15% |  |
| Freezing of gait reported at visit | - | 37% |  |
| Age at onset (years) | - | 60.1 ± 10.3 |  |
| Disease duration (years) | - | 9.2 ± 5.7 |  |
| Hoehn & Yahr score | - | 2.1 ± 0.8 |  |
| Daily levodopa dose (mg/day) | - | 667 ± 368 (n=23) |  |
| levodopa per dose (mg/dose) | - | 194 ± 84 (n=23) |  |
| Duration on levodopa (years) | - | 5.3 ± 3.2 (n=23) |  |
| On agonist at visit | - | 7% |  |
| On MAO-I at visit | - | 33% |  |
| LEDD (l-dopa+agonist+MAO-I) | - | 695 ± 393 (n=24) |  |

### Gender and disease asymmetry of finger tapping spatiotemporal parameters

The output of the data gloves provides a sinusoidal trace of finger tapping as shown in Figure 1A for a PD participant. Tap amplitude (trough to peak, tap amplitude) and intertap interval (peak-to-peak time, illustrated as tap interval) measures are illustrated in Figure 1A. Additional sample traces of control participants and PD participants both OFF and ON levodopa are shown in Supplementary Figure 1. As the gender distribution was different between the PD and control groups (Table 1), we explored finger tapping kinematics by gender in each group. No significant differences in any measure of upper limb kinematics was found between genders in either group (Table 2), so measures were collapsed across gender.

**Table 2:** Gender grouped upper limb kinematics

|  | controls |  |  | PD |  |  |
| --- | --- | --- | --- | --- | --- | --- |
|  | female (n=16) | male (n=8) | p-value | female (n=8) | male (n=19) | p-value |
| mean tap amplitude (norm. units) | 164.49 ± 46.09 | 182.27 ± 27.33 | 0.250 | 132.97 ± 56.41 | 139.36 ± 51.27 | 0.787 |
| CV tap amplitude (%) | 20.06 ± 12.02 | 17.48 ± 13.22 | 0.651 | 20.06 ± 12.02 | 17.48 ± 13.22 | 0.651 |
| mean tap interval (s) | 0.76 ± 0.25 | 1.05 ± 0.47 | 0.140 | 0.65 ± 0.27 | 0.67 ± 0.28 | 0.883 |
| CV tap interval (%) | 18.70 ± 12.69 | 18.70 ± 16.97 | 0.999 | 24.39 ± 18.36 | 25.37 ± 17.91 | 0.901 |
| tap frequency (taps/s) | 1.40 ± 0.50 | 1.08 ± 0.58 | 0.211 | 1.64 ± 0.56 | 1.83 ± 1.09 | 0.559 |
| All values reported as mean ± standard deviation |  |  |  |  |  |  |

Baseline upper limb kinematic features between control and PD participants in the OFF levodopa state (PD-OFF) are illustrated in Figure 1. At a self-defined normal tapping speed, there were significant differences in the more affected (MA) but not less affected (LA) hand in PD-OFF compared to control participants. For amplitude measures in the MA hand, PD-OFF had a smaller mean tap amplitude (Figure 1D) and greater CV tap amplitude compared to controls (Figure 1E). For our secondary comparisons, mean tap interval was also shorter (Figure 1B), and tap frequency faster (Figure 1F) in PD-OFF compared to controls. PD participants tended to show more tachykinetic (smaller amplitude, faster tapping) finger tapping than controls. As differences were only observed in the MA hand during the bimanual tapping task, all subsequent analysis is shown for the MA hand only.

### Comparison of finger tapping and gait spatiotemporal parameters

To determine the relationship between upper limb and gait kinematics, we examined finger tapping and gait in the MA hand and MA leg in PD-OFF participants. Figure 2 shows regression plots for mean (Figure 2A) and CV (Figure 2D) tap amplitude compared to stride length in PD-OFF (gray circles) and controls (black diamonds) as equivalent amplitude measures. We found a significant positive correlation between mean tap amplitude and stride length (Pearson’s= 0.418, R^2^=0.174, p=0.030) in PD-OFF. Secondary analysis was performed comparing tap amplitude to other gait measures, UPDRS and MoCA scores (Table 3). In PD-OFF participants tap amplitude was inversely correlated with motor and total UPDRS scores and also with the bradykinesia subscores of the motor (sum of UPDRS items 23-26 and 31) and ADL sections (sum of UPDRS items 9-12), even after adjustment for multiple comparisons using a Benjamini-Hochberg adjustment.

**Figure 2:**
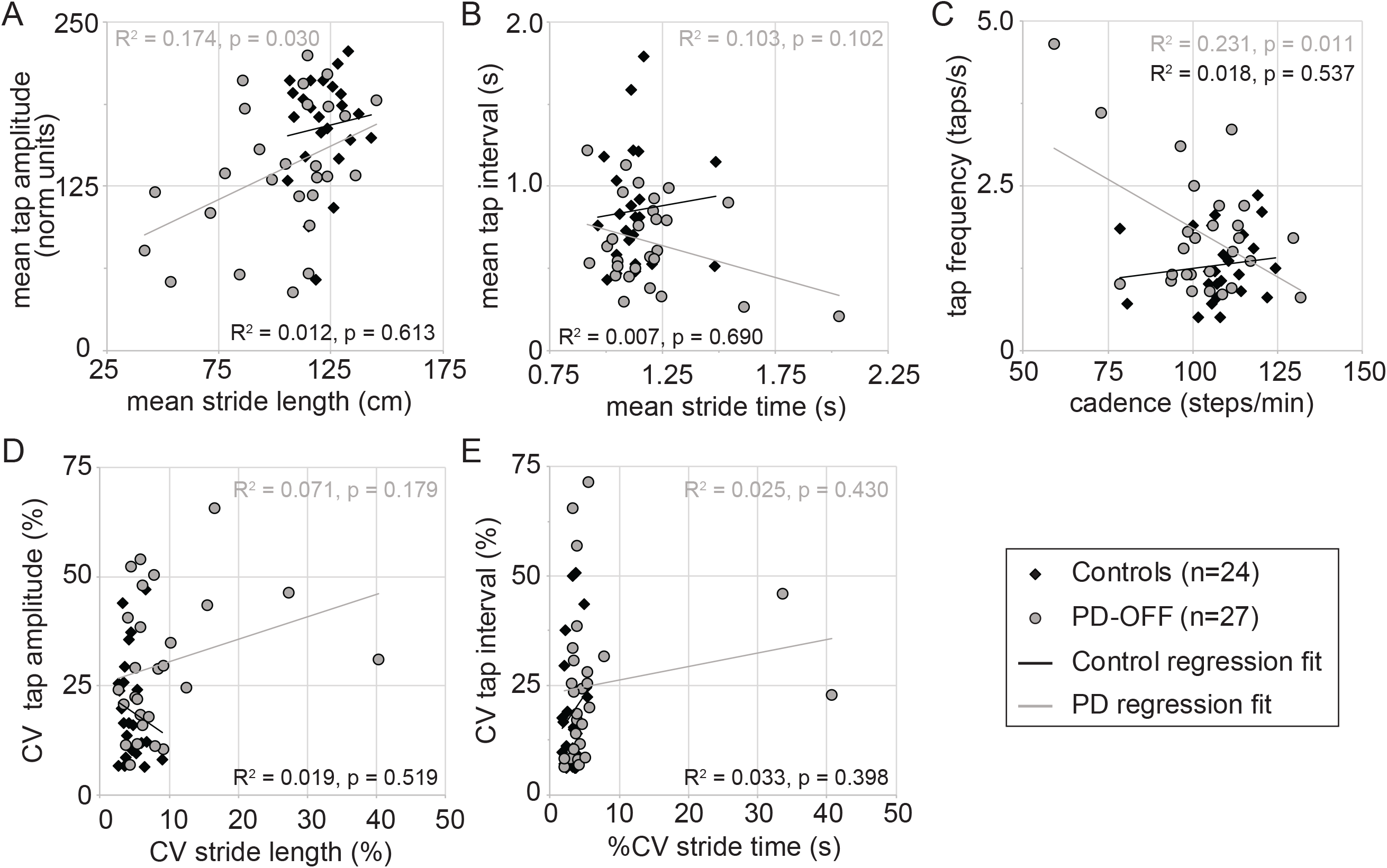
Spatiotemporal parameters of upper limb finger tapping and gait for the more affected limb. Mean and variability (CV) measures for finger tap amplitude vs stride length **(A, D)** finger tap interval vs stride time **(B, E)**, and tap frequency vs cadence **(C)** for self-defined normal “comfortable” paced movements of the dominant limb are shown for controls (black diamonds) and PD participants OFF levodopa (PD-OFF). Each symbol represents a single participant. Best fit lines for PD and control participants are also shown.

**Table 3:**
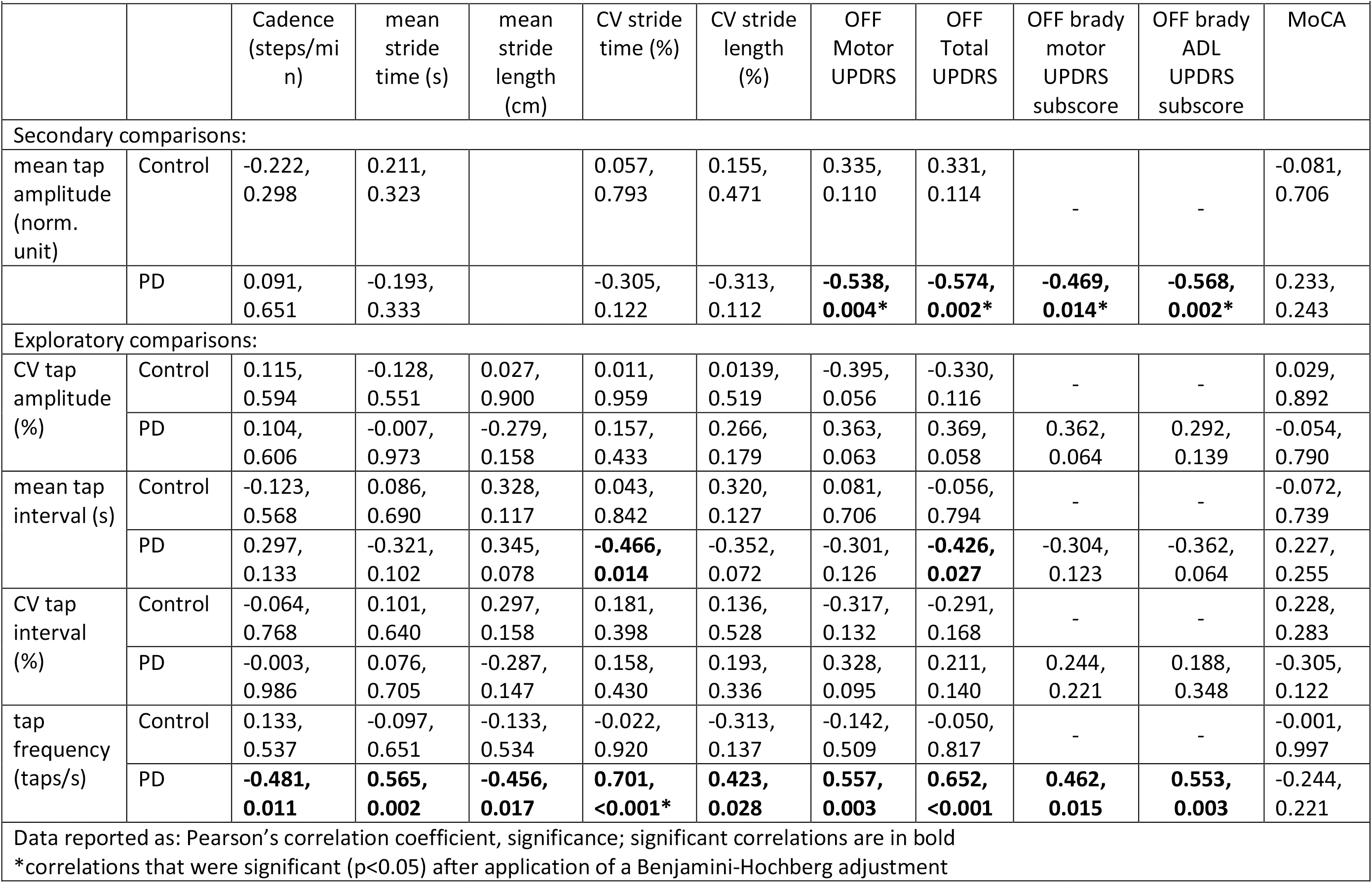
Pearson’s correlation coefficients comparing disease dominantly affected upper limb kinematics, gait and UPDRS scores

Exploratory analysis was undertaken to compare the finger tapping and gait measures of timing (Figure 2B, E) and frequency (Figure 2C) of movement. In PD-OFF there was a negative correlation between tap frequency and cadence (Pearson’s=-0.418, R^2^=0.231, p=0.011). In PD-OFF participants, tap frequency was also found to be correlated to all gait measures (Table 3) and remained significantly correlated with mean stride length, CV stride length, motor, total and bradykinesia ADL UPDRS scores after adjusting for multiple comparisons. No significant correlations were found in in control participants.

### Finger tapping manipulating motor-load

In order to manipulate motor load, participants were asked to tap at a self-defined fast speed. Finger tapping was not entrained to a particular metronome frequency, as typically gait assessments are also not set to any particular frequency and we did not want to constrain variability in the responses. In both PD-OFF and controls, there was a smaller mean tap amplitude (Figure 3A) in the fast compared to normal speed. Using a 2×2 multifactorial ANOVA with speed (fast-slow) and group (PD-OFF/controls) as factors there was a between-subject main effect for tap amplitude (p=0.018) but no within-subject effect for speed × group. Tap amplitude CV was higher in PD-OFF compared to control at both speeds (Figure 3B) but there was not a statistically significant between-subject main effect (p=0.018) or a within-subject effect for speed × group.

**Figure 3:**
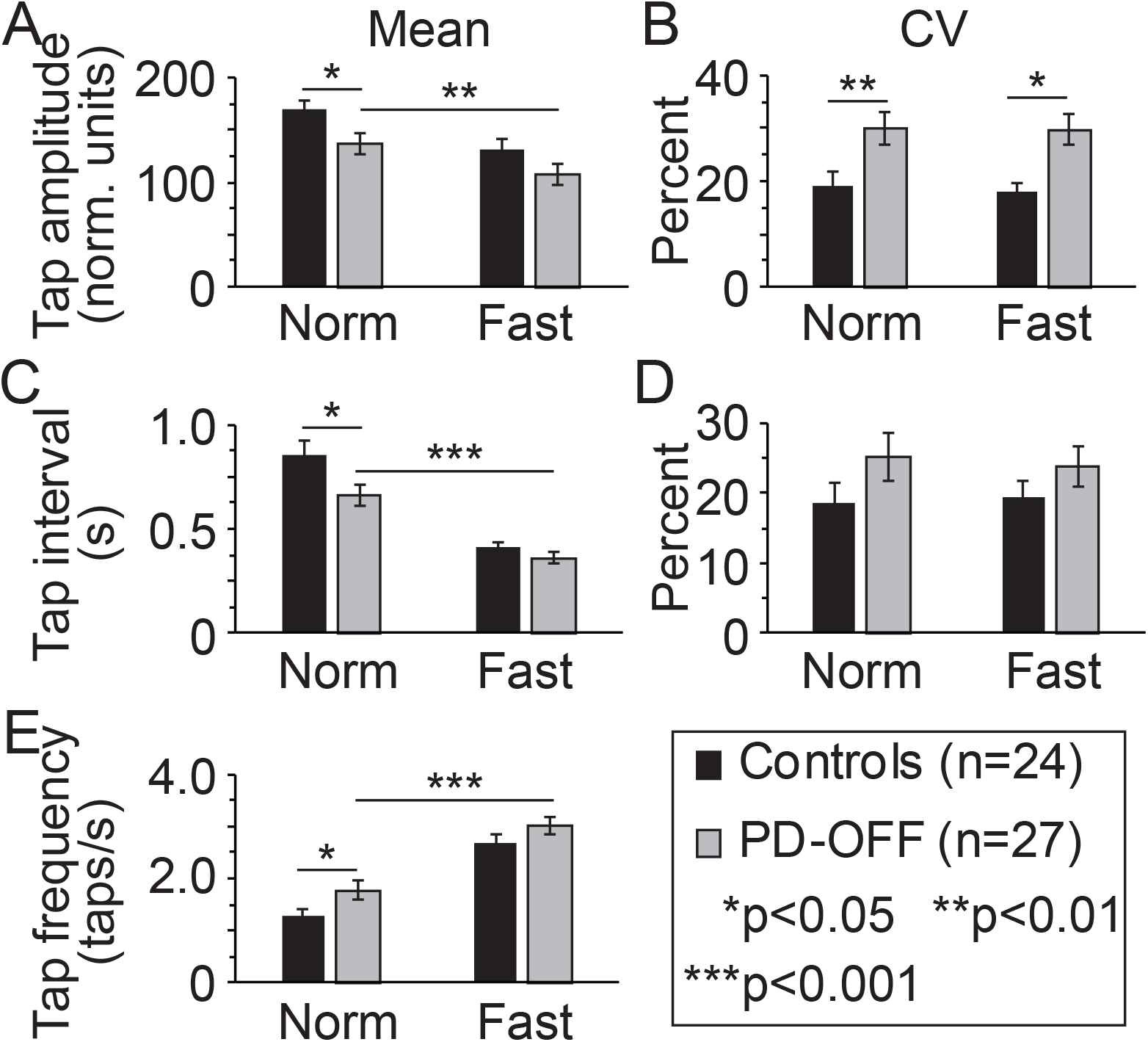
Spatiotemporal parameters of upper limb finger tapping manipulating tap speed in the more affected hand. Mean and variability (CV) in finger tap amplitude **(A, B)** finger tap interval **(C, D)**, and tap frequency **(E)** for self-defined normal (Norm) and fast paced (Fast) finger tapping are shown for controls and PD participants OFF levodopa (PD-OFF). Bar graphs are plotted as the mean ± sem.

On secondary measures, PD-OFF had a shorter mean tap interval (Figure 3A) and faster tap frequency (Figure 3E) at the faster tap speed. There were between-subject main effects for mean tap interval (p=0.033) and tap speed (p=0.034), but not CV tap interval (p=0.139). There were no within-subject effects for speed × group.

### Levodopa response in finger tapping and gait spatiotemporal parameters

The dynamics of the levodopa response in the MA hand and leg in 21 PD participants is shown in Figure 4. Six PD-OFF participants were not included in this analysis due to the following: 4 were not clinically treated with levodopa for symptomatic management, 1 participant forgot to bring levodopa to the visit, and 1 participant’s ON-levodopa finger tapping data was corrupted and could not be used. Mean tap amplitude and mean stride length significantly increased at faster speed (Figure 4A, F) in the ON-levodopa state (PD-ON). At normal speed, although both increased in the ON-state, the increase did not reach statistical significance for tap amplitude (p=0.129). There was also a trend for CV stride length and tap amplitude to decrease in the ON-state (Figure 4 B, G), but statistical significance was present only at normal speed. ON secondary measures, while tap frequency was faster (Figure 4E) and tap interval shorter (Figure 4C) in the ON-state fast speed, cadence (Figure 4J) and stride time (Figure 4I) were not significantly different.

**Figure 4:**
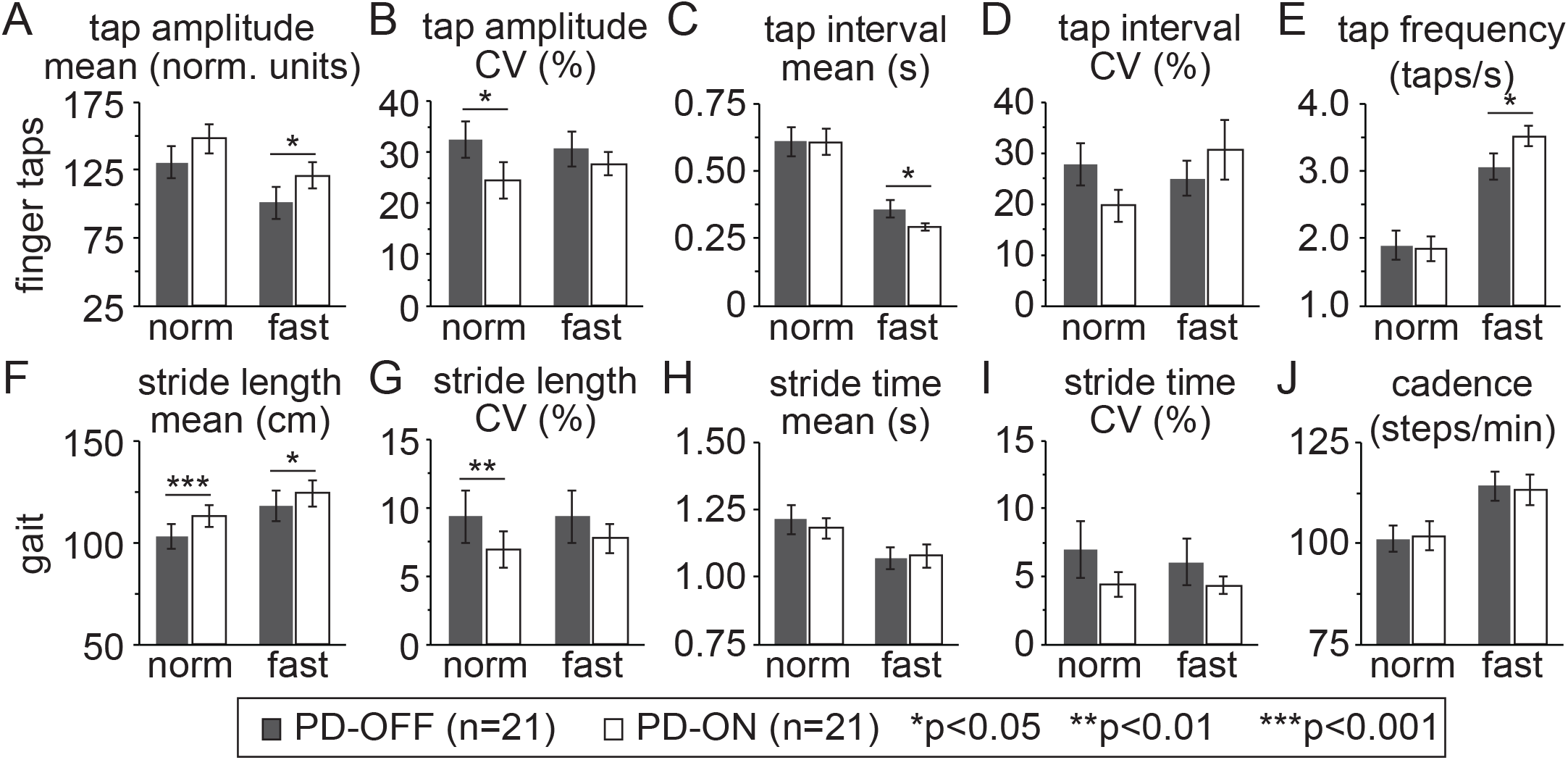
Levodopa responsiveness of spatiotemporal parameters of upper limb finger tapping and gait. Levodopa responsiveness in upper limb spatiotemporal parameters of mean and variability (CV) in finger tap amplitude **(A, B)**, finger tap interval **(C, D)**, and tap frequency **(E)** are shown in the top row for PD participants OFF levodopa (PD-OFF) and ON levodopa (PD-ON). Levodopa responsiveness in gait spatiotemporal parameters of mean and variability (CV) in stride length **(F, G)** stride time **(H, I)**, and cadence **(J)**, are shown in the bottom row. Bar graphs are plotted as the mean ± sem.

We also calculated the levodopa response as the difference between the OFF and ON result (delta) for each participant for each gait parameter, tapping parameter and the motor UPDRS at normal speed. The delta tap amplitude and delta stride length were correlated with each other (Pearson’s=0.484, R^2^=0.235, p=0.026) (Figure 5A). The delta motor UPDRS was also inversely correlated with the delta tap amplitude (Figure 5B) (Pearson’s=-0.605, R^2^=0.336, p=0.004) and delta stride length (Figure 5C) (Pearson’s=-0.441, R^2^=0.194, p=0.046). These were significant even adjusting for the multiple comparisons.

**Figure 5:**
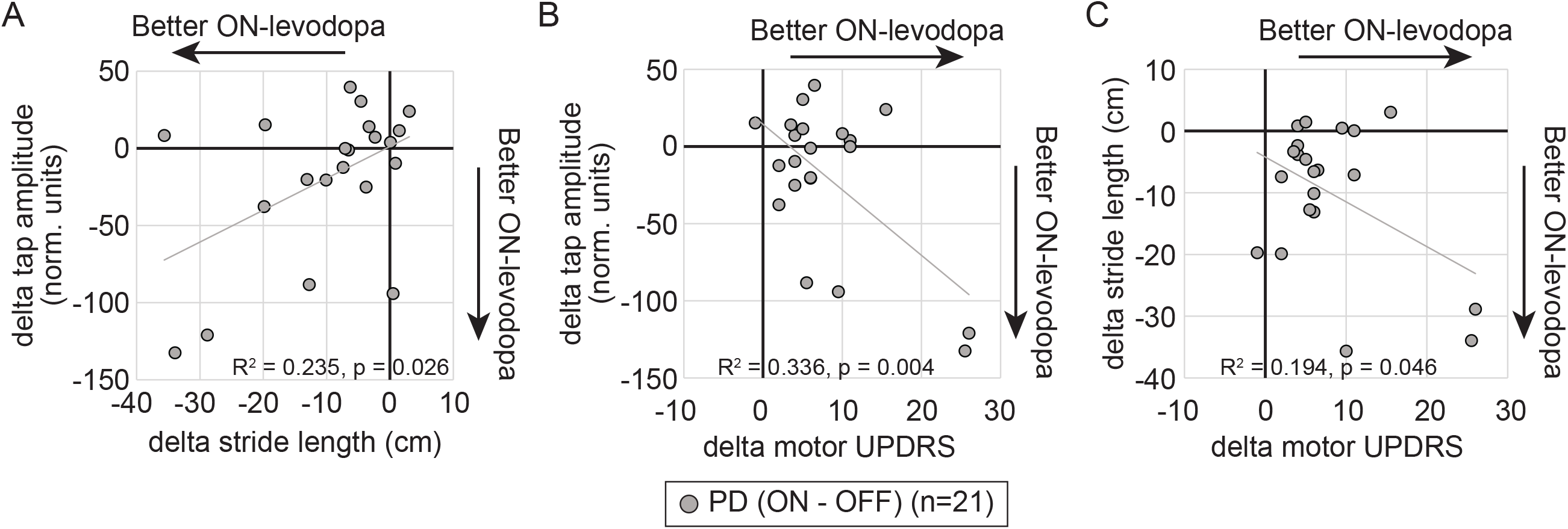
Comparison of levodopa response in tapping amplitude, gait and motor UPDRS scores in participants with Parkinson’s disease. Mean change in amplitude measures of finger tapping, gait stride length and motor UPDRS scores between the ON and OFF conditions are shown for **(A)** tap amplitude vs stride length, **(B)** tap amplitude vs motor UPDRS scores and **(C)** stride length and motor UPDRS scores. Each symbol represents a single participant. Best fit lines are also shown.

## Discussion

Amplitude setting is impaired in PD, whether it be in the form of bradykinetic finger or foot tapping, micrographic handwriting, or decreased stride length when walking. One of the primary findings of our study was that amplitude setting of the more disease affected side during bimanual anti-phase tasks in the upper limb and lower limb were correlated and showed a similar response to motor load and to levodopa. This finding is important for multiple reasons. Firstly, clinically PD patients are often divided into two groups: 1) upper body or tremor predominant Parkinsonism, with more tremor and/or upper limb bradykinesia being predominant features, or 2) lower body Parkinsonism with more shuffling gait and postural instability. Since our study suggests that the severity of amplitude deficits are comparable in both upper and lower limbs in individuals, focusing on the predominant disease feature while planning treatments may limit overall functional improvement. Secondly, from a research standpoint, pathways for modulation of gait are difficult to study; functional magnetic resonance imaging (fMRI) cannot be performed while walking. However, our finding that the response to motor load and levodopa affects amplitude setting similarly both in finger tapping and gait, suggests that upper limb bimanual movements could be used as a simpler model system when studying this important feature of PD dysfunction.

While gait control also involves balance control circuity that would not be activated in a finger tapping task, our finding that smaller tap amplitudes were correlated in the same PD participants with shorter stride length, suggest that there are underlying circuits that modulate amplitude during tasks requiring bilateral coordinated movement. Levodopa improved both tap amplitude and stride length, at least in the fast conditions, suggesting that, at least in part, dopaminergic pathways may be involved. Stride length is shorter in PD (Morris et al. 1996), and a sequential decrease in stride length (termed the sequence effect) has been suggested as a mechanism for freezing of gait episodes (Chee et al. 2009; Virmani et al. 2018). As in other studies (Nieuwboer et al. 2009; Stegemoller et al. 2009), we find that PD participants, at least in the MA hand, have smaller tap amplitudes. Smaller amplitude movements have been used to elicit freezing in an upper limb bimanual tapping task (Williams et al. 2013) as well is handwriting (Heremans et al. 2015). Whether a similar sequence effect leads to upper limb freezing episodes as it does in gait remains to be seen.

Besides absolute amplitude setting, variability or arrhythmicity in movement amplitude between successive repetitive movements is also important in PD and is measured as the percent coefficient of variation in amplitude across a trial. In our study we report an increase in tap amplitude variability in the MA (but not LA) hand in PD compared to control, similar to that seen using a MIDI keyboard tapping task (Trager et al. 2015). Amplitude variability has also been correlated to the presence and duration of upper limb freezing episodes (Barbe et al. 2014). While we did not find a correlation between the CV in finger tapping amplitude and gait stride length CV in PD-OFF, both improved in the levodopa ON-state (PD-ON) compared to OFF-state (PD-OFF) condition, suggesting that upper limb and gait rhythmicity may be modulated through common dopaminergic pathways as well.

PD is an asymmetric disease, usually starting on one side, and maintaining asymmetry even once the other side is involved. In our study, we were able to objectively measure the asymmetry in PD upper limb function when compared with control participants. Consistent with our findings, in a prior study using a MIDI keyboard, decreased velocity of tapping in the more affected hand was detected in untreated PD participants within 1.5 years of diagnosis (Koop et al. 2008). Dual-task behaviors have also been shown to preferentially affect performance in the more affected compared to less affected hand (Acaroz Candan and Ozcan 2019). Compared to gait freezing, which typically occurs in both legs, unilateral freezing in upper limb tasks has been reported (Vercruysse et al. 2012a). Asymmetry index calculations often utilize right and left limbs (Plotnik et al. 2008) not more and less affected limbs so care must be taken when interpreting results, especially since these studies have suggested gait asymmetry as a cause of freezing.

Whether levodopa affects upper limb movement similarly to gait is also unclear from past studies. In our study, with upper limb movement and gait measurements made in the same people on the same day, we found that in the PD-ON condition participants tapped with a larger amplitude and walked with a longer stride length compared to PD-OFF, especially in the fast speed condition. Variability in finger tap amplitude and stride length was also reduced in PD-ON compared to the PD-OFF state. In prior studies of upper limb repetitive movements, levodopa has been shown to have mixed results. Amplitude measures have been shown to improve (Almeida and Brown 2013) or remain unchanged with levodopa (Barbe et al. 2014; Stegemoller et al. 2009). Amplitude variability in gait freezers but not non-freezers has been shown to improve (Barbe et al. 2014). Tap speed has been shown to improve (Taylor Tavares et al. 2005). Tap interval was decreased by levodopa in one study (Taylor Tavares et al. 2005) but had no effect in others (Barbe et al. 2014; Stegemoller et al. 2009). Upper limb freezing frequency also possibly decreased (Barbe et al. 2014) or remained unchanged (Brown et al. 2015). Barbe and colleagues (Barbe et al. 2014) also show levodopa responsiveness in stride length in their participants but unlike in our study, they do not directly compare these responses in the upper limb and gait measure.

The common pathways affecting both upper and lower limb amplitude setting are not clearly defined as yet. Studies focused on either upper limb movements or imagined gait have found differing areas of altered brain activation or connectivity. This could be due to different upper limb maneuvers used in different studies in addition to low numbers of participants. However, a recent exploratory meta-analysis suggested that the cerebellar locomotor region showed the most consistent gait-related activation in PD (Gilat et al. 2019). Two finger tapping studies, one using a sequential finger tapping task (Wu and Hallett 2005), and another using a motor timing task also showed greater activation of the bilateral cerebellum (Jahanshahi et al. 2010). This suggests that cerebellar connectivity may be an important area to explore in future studies of common pathways of upper and lower limb dysfunction. Based on our results, an upper limb task-based paradigm using dynamic fMRI, while manipulating the amplitude of finger tapping might help define network level changes responsible for impaired amplitude setting common to gait and finger tapping. This could in turn help develop therapies to treat levodopa resistant bradykinesia/tachykinesia and gait deficits.

Limitations of this study include that fact that a single size data glove was utilized so we cannot exclude the possibility that people with larger hands might have had suboptimal sensor positioning leading to some of the observed variability. However, for most participants the gloves fit snuggly and this should be distributed equally amongst the control and PD groups. Additionally, gender distribution differed between the PD and control groups and could affect our comparisons. However, on a subgroup analysis (Table 2) we did not see significant gender based differences in our results in either group. As there are a number of parameters tested (amplitude, timing, cadence), under different conditions (fast/normal speed, ON/OFF levodopa), we understand that we may be overestimating the number of variables of interest that were statistically significantly different from one another. Our study was also not focused on a comparison of methods for determination of upper limb kinematic measures. However, we note that analogue encoders placed on the rotation axis of the index finger have previously been used to objectively define finger tapping movements in bimanual finger tapping studies (Vercruysse et al. 2012b; Vercruysse et al. 2014). The data glove provided sinusoidal traces from which amplitude and timing measures could be calculated, similar to gait measures of stride length and time obtained from the gait mat. The resultant traces showed decreased amplitude in finger tapping in PD compared to controls and levodopa responsiveness in PD participants as we would clinically expect. Additionally, upper limb spatiotemporal parameters of tap frequency and tap amplitude were correlated with total motor UPDRS scores and sub-scores of bradykinesia. Taken together these findings suggest that our data glove measurements are reflecting disease pathology.

## Conclusions

In summary, we show that upper limb and gait amplitude measures were correlated, and showed similar dopaminergic response and response to motor loading. This suggests these automated motor functions are subserved by common functional networks. Future studies utilizing tasks manipulating upper limb amplitude during dynamic fMRI could help provide greater understanding of the networks that modulate movement amplitude, leading eventually to substrates for future therapeutic intervention.

## Data Availability

Anonymized data sets can be shared at the request of qualified investigators by direct correspondence.

## Acknowledgments

This work was supported in part by the University of Arkansas Clinician Scientist Program (UAMS CSP)(TV), the University of Arkansas Development Enhancement Award Program (UAMS DEAP) (LLP), and the Arkansas Children’s Nutrition Center (USDA 6026-51000-012-06S)(LLP). We also greatly appreciate the commitment and dedication of our participants, without whom this work would not be possible.

## Declarations

### Ethics approval

The study was approved by the University of Arkansas for Medical Sciences institutional review board (UAMS IRB# 228861) and the study was conducted in accordance with the guidelines of the Declaration of Helsinki.

### Consent to participate

Written informed consent was obtained from all participants prior to performing any study assessments.

### Consent for publication

All authors have reviewed the manuscript and agree to publication in its current form.

### Code Availability

The peak and trough detection code can be shared at the request of qualified investigators by direct correspondence.

### Author contributions

Lakshmi Pillai, Aliyah Glover, Aaron Kemp and Hafsa Syeda were involved in research project organization and execution and manuscript review and critique. Horace Spencer was involved in statistical design and execution and manuscript review and critique. Linda Larson-Prior was involved in research project conception, organization, statistical design, and critique and revision of the manuscript. Tuhin Virmani was involved in research project conception, organization and execution, statistical design and execution, and writing and revision of the manuscript.

---

**Supplementary Figure 1:**
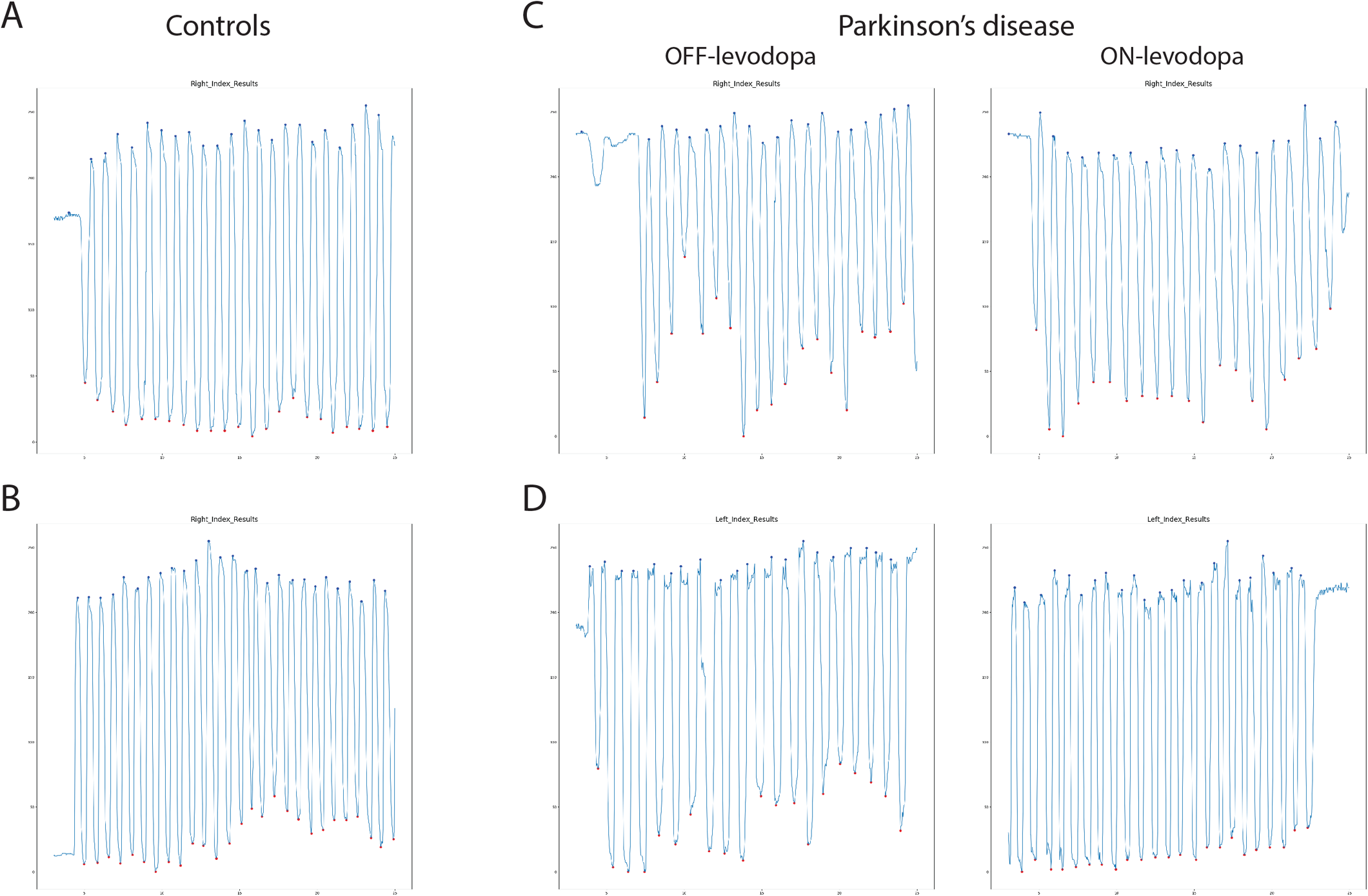
Sample traces of finger tapping raw output from the 5DT data glove with red and blue dots from our algorithm defined peaks and troughs from which spatiotemporal measures were calculated. **(A, B)** raw output from 2 control subjects dominant hand are shown. **(C, D)** raw output from 2 PD subjects more affected hands showing smaller amplitudes and greater amplitude variability in the OFF-levodopa condition (left panel) compared to the ON-levodopa condition (right panel).

